# Contaminating manufacturing plasmids and disrupted vector genomes present in liver tissue following adeno-associated virus gene therapy

**DOI:** 10.1101/2025.01.13.25320105

**Authors:** Sarah Buddle, Li-An K Brown, Sofia Morfopoulou, Oscar Enrique Torres Montaguth, Mariacristina Scoto, Vanessa Herder, Anil Dhawan, Julianne R Brown, Laura Atkinson, Angelika Kopec, Dee Davis, Nathaniel Storey, Luis Campos, Neil Sebire, Hannah Macpherson, Jasmaine Lee, Richard Orton, Giovanni Baranello, Patawee Asamaphan, Georgios Ilia, Rajvinder Karda, Holly Belfield, ISARIC 4C Investigators, Malcolm Gracie Semple, J Kenneth Baillie, John Counsell, Simon Waddington, Emma C Thomson, Francesco Muntoni, Judith Breuer

**Author notes:** These authors contributed equally to this work (Sarah Buddle, Li-An K Brown). See consortium list (ISARIC 4C Investigators).

## Abstract

Adeno-associated viruses (AAVs) are the most used vectors in gene therapy but can frequently cause liver complications in patients. The mechanisms underlying AAV-related liver toxicity remain poorly understood, posing challenges for effective prevention and intervention. We undertook long and short read metagenomic sequencing of liver tissue from a child with spinal muscular atrophy type 1 experiencing significant hepatitis after receiving onasemnogene abeparvovec. We identified manufacturing plasmid sequences, with evidence of complex structures and recombination. Vector genomes had extensive disruption and concatemerisation. We also identified the presence of human betaherpesvirus 6B in the liver. It is possible that presence of the manufacturing plasmid sequences or helper viruses allow replication of the vector within cells, contributing to the development of complex concatemeric structures and associated hepatitis.

## Main

AAV gene therapies show promise for treating a wide variety of serious genetic conditions, such as haemophilia^1–3^, muscular dystrophies^4^ and spinal muscular atrophy^5^. As of 2024, there were seven AAV gene therapies approved by the FDA^6^, with many more in clinical trials. The most common adverse effect of intravenously-administered AAV gene therapies is hepatotoxicity, with most patients experiencing a rise in serum liver enzymes, which is routinely treated with high dose steroids. Occasionally, liver toxicity is severe and some patients have experienced fulminant liver failure^7–10^. Hepatotoxicity tends to be more severe in older patients with a higher body weight, who receive higher vector doses, and who may require additional immunosuppression^11,12^. Pre-existing liver disease appears to be a significant risk factor^7^.

The mechanisms underlying hepatotoxicity are complex and incompletely understood. Hepatitis has been postulated to be caused by innate, humoral and cellular immune responses to the vector capsid, genome, or transgene product^13–15^, or by impurities within the vector preparation^16,17^, or from a direct toxic effect^18,19^. Acute sinusoidal endothelial injury resembling capillary leak syndrome has also been well documented in non-human primates using both empty capsids and therapeutic transgenes^20^.

Onasemnogene abeparvovec (Zolgensma®, OA) is an AAV-vectored gene therapy for spinal muscular atrophy (SMA), a neurodegenerative disease caused by deleterious variants in the *SMN1* gene which encodes the survival motor neuron protein (SMN)^21^. OA is manufactured using three plasmids (**Figure 1**): the vector plasmid (pSMN), which contains *SMN* and a promotor region between two AAV inverted terminal repeats (ITRs), pAAV2/9, which contains AAV2 *rep* and AAV9 *cap* genes, and pHelper, which contains the human adenovirus (HAdV) genes necessary for AAV replication^22,23^. The resultant vector preparation contains therapeutic recombinant AAV (rAAV) particles which have an outer AAV9 capsid, containing a human *SMN* coding sequence between AAV ITRs, in a self-complementary structure. Apart from this desirable construct, there are also manufacturing process-related impurities including empty capsids, reverse packaged manufacturing plasmids, genome fragments, and recombined products^24,25^. Transcriptionally-active sequences from a Rep-Cap manufacturing plasmid have been identified in mouse liver after systemic administration of a good manufacturing practice (GMP) produced and purified rAAV preparation^25^. These manufacturing issues are complex to study and resolve, and the FDA has released guidance on reporting and validating the steps in the manufacturing process^26^.

**Figure 1:**
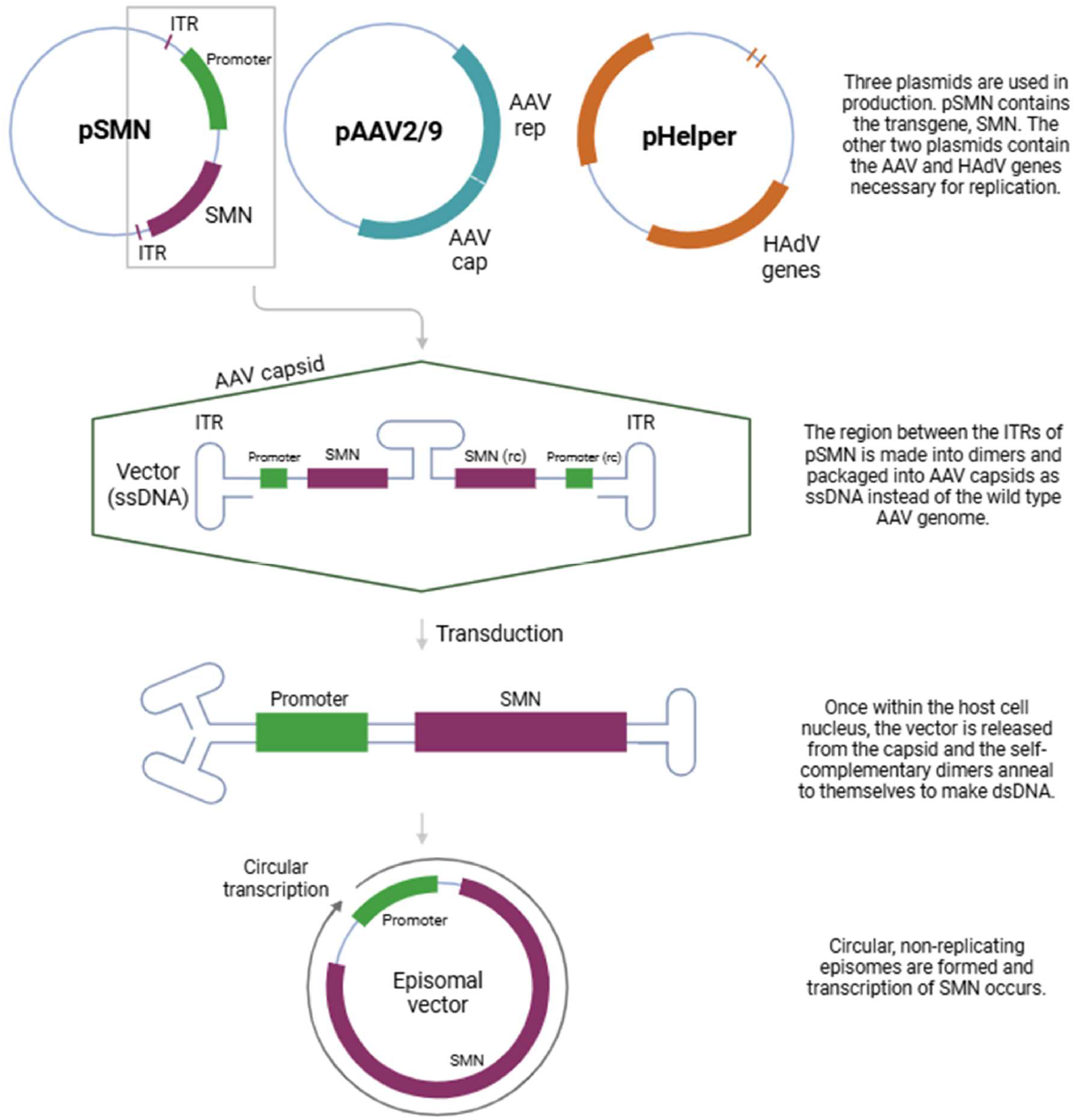
Schematic of plasmids used to manufacture onasemnogene abeparvovec and its mechanism of action. OA is produced by transfection of HEK293 cells with a vector plasmid (pSMN), containing *SMN* between AAV inverted terminal repeats (ITRs), an AAV plasmid containing AAV2 *rep* and AAV9 *cap* genes (pAAV2/9), and a helper plasmid containing HAdV genes such as E2A, E4 and VA RNA genes (pHelper)^22,23^. SMN, survival motor neuron; AAV, adeno-associated virus; HAdV, human adenovirus; ITR, inverted terminal repeat; ssDNA, single-stranded DNA; dsDNA, double-stranded DNA. Produced using biorender.com.

We investigated a patient treated with OA for type 1 SMA who experienced significant symptomatic hepatitis after infusion. [*Potentially identifying clinical details withheld in preprint*]

Histology of the liver biopsy showed a marked periportal and lobular lymphocytic infiltrate with interface inflammation, patchy hepatocyte necrosis and ballooning degeneration, with relative sparing of bile ducts (**Figure 2A, B**). This histological pattern is similar to that seen in children with hepatitis associated with wild type AAV2 infection^27,28^ and in “indeterminate” paediatric acute liver failure^29^. *In situ-*hybridisation for *SMN* confirmed hepatocyte vector transduction in the patient liver (**Figure 2C, D**).

**Figure 2:**
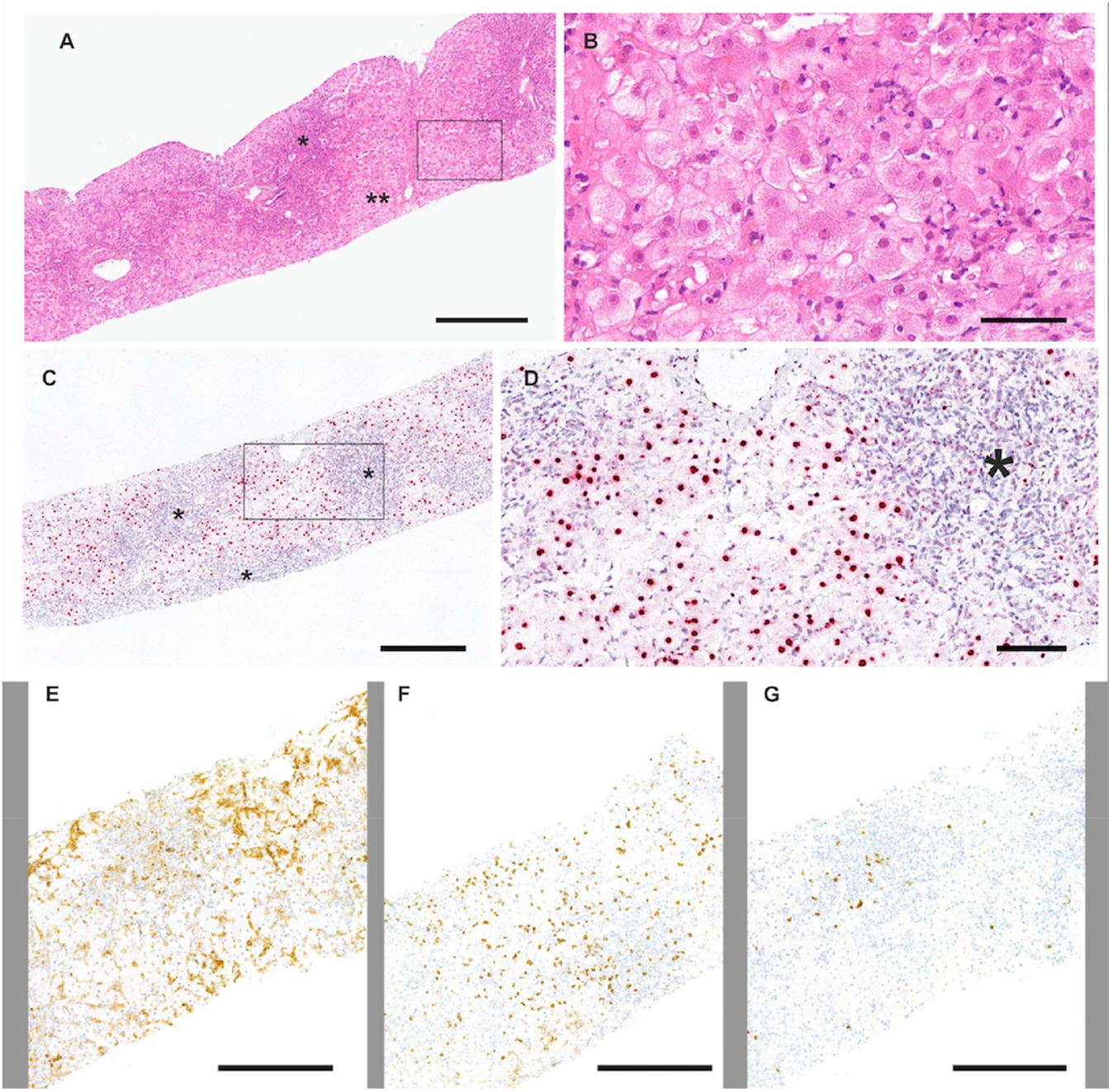
Liver biopsy findings. A) Liver biopsy of the patient shows marked periportal and lobular inflammation as well as interface inflammation (^*^); numerous hepatocytes with ballooning degeneration are present (^**^). Box in A is magnified in B. B) High magnification of ballooning hepatocytes highlighting the swollen cytoplasm. C) RNA in situ-hybridisation detecting the SMN1-gene shows a strong positive red signal in the nucleus of ballooning hepatocytes separated by areas with severe immune cell infiltration (^*^). Box in C is magnified in D. D) High magnification of ballooning hepatocytes shows the positive signal in the nucleus and a mild to moderate, punctuated signal within the cytoplasm of hepatocytes and of the immune cells (^*^). Inflammation in the liver is shown by immunohistochemistry detecting CD4 (E), CD8 (F) and CD20 (G). Bars, A and C, 400 micrometres, B, 60 micrometres, D, 100 micrometres, E-G, 300 micrometres.

We conducted untargeted short-read metagenomic sequencing of DNA and RNA from the residual patient liver sample. Analysis identified multiple serotypes of AAV, human mastadenovirus C (HAdV-C) and human betaherpesvirus 6 type B (HHV-6B) in the DNA-seq, while RNA-seq did not identify any pathogen transcripts, consistent with lack of viral replication (**Table 1**). For AAV2 and HAdV-C, the sequencing reads only aligned to the sections of the viral genomes that are part of the OA manufacturing plasmids, suggesting presence of plasmids in the liver tissue rather than wild-type virus infection (**Figure 3A**). In contrast, HHV-6B reads covered the breadth of the HHV-6B genome (**Figure 3B**), suggesting natural HHV-6B infection. Accordingly, a specific PCR for HAdV was negative (targeting a region of the genome which is not present in pHelper), and a specific PCR for HHV6 was positive (Ct value= 26.2). Aligning reads to the manufacturing plasmid sequences, we found good coverage of the vector genome, but also of the other regions of the pSMN manufacturing plasmid and of pAAV2/9, with some reads mapping to pHelper (**Figure 3C**).

**Table 1.**
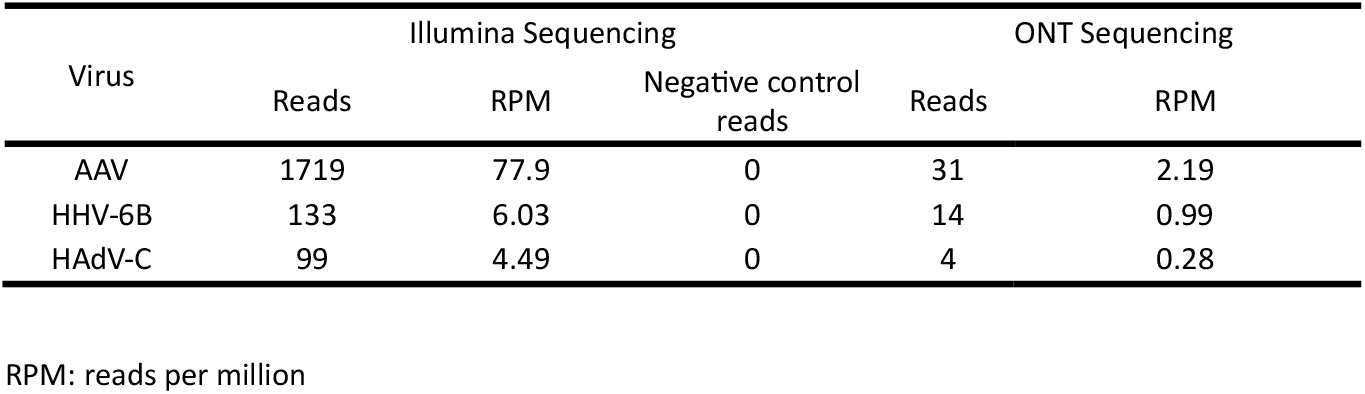
Metagenomic sequencing results.

| Virus | Illumina Sequencing |  |  | ONT Sequencing |  |
| --- | --- | --- | --- | --- | --- |
|  | Reads | RPM | Negative control reads | Reads | RPM |
| AAV | 1719 | 77.9 | 0 | 31 | 2.19 |
| HHV-6B | 133 | 6.03 | 0 | 14 | 0.99 |
| HAdV-C | 99 | 4.49 | 0 | 4 | 0.28 |

RPM: reads per million

**Figure 3:**
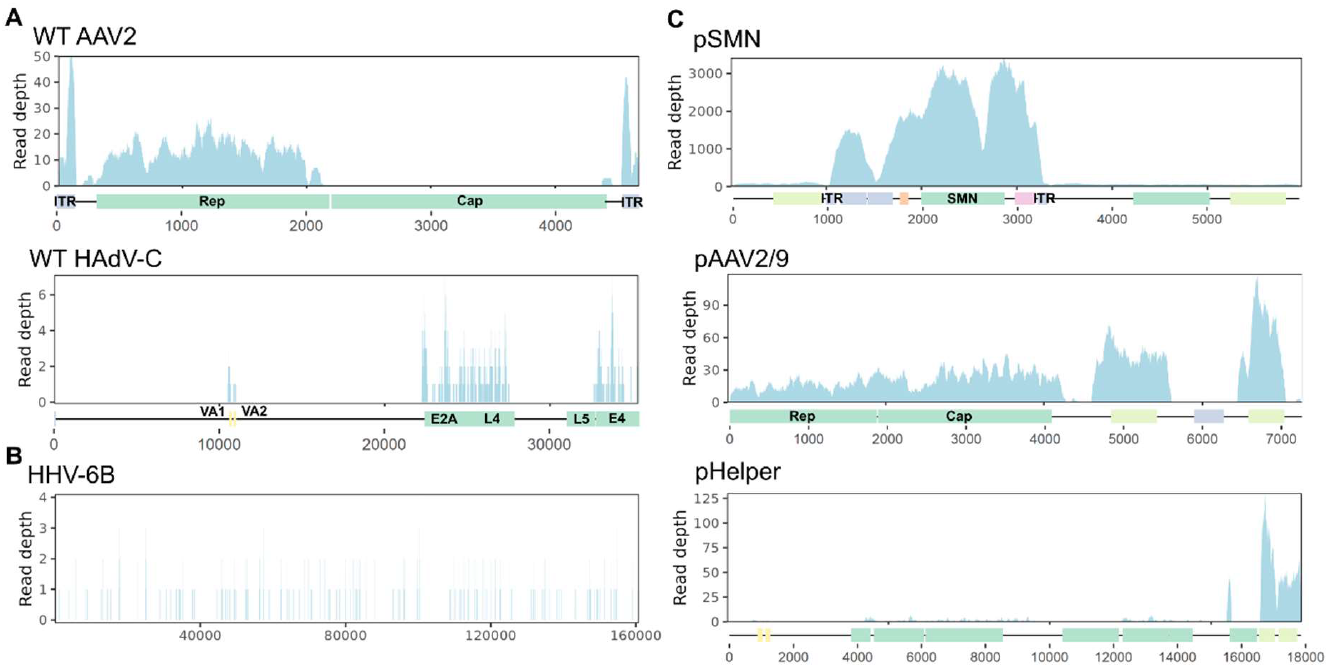
Metagenomic sequence alignment. A) Genome coverage of wild-type (WT) AAV2 and HAdV-C from Illumina sequencing reads. Approximate locations of the genes present in the manufacturing plasmids are marked along the x-axis. AAV2 alignment uses more stringent mapping parameters to more clearly differentiate between any AAV2 and AAV9 derived sequences – see methods. B) Alignment of Illumina sequencing reads to the HHV-6B genome shows reads cover the breadth of the genome. C) Alignment of Illumina sequencing reads to approximate manufacturing plasmid sequences shows presence of plasmid sequences. NB From the negative control, 10 reads aligned to the pSMN sequence. No reads aligned to the pAAV2/9 or pHelper sequences.

To further confirm the presence of the manufacturing plasmids in the liver, we performed untargeted long read metagenomic sequencing of the liver sample (Oxford Nanopore Technologies). Again, we found reads aligning to AAVs, HHV-6B and HAdV-C (**Table 1**), along with the gene therapy vector sequence. Reads mapping to all three of the manufacturing plasmids were also found, primarily to pSMN (including the region of the plasmid that is not included in the therapeutic vector genome), and pAAV2/9 (**Table 2**). Sequence analysis of individual reads showed high levels of vector genome concatemerisation and complex genome structures with rearrangements (**Figure 4A-D, Table 2**). The concatemeric patterns observed show similarities to those seen in replicating AAVs using rolling hairpin and rolling circle amplification^30^. Many of the structures observed were longer than the maximum packaging length of an AAV vector (approximately 4.7kb), suggesting that recombination may have occurred *in vivo*. Alternatively, these sequences may represent plasmid contaminants from manufacture. The majority of pAAV2/9 reads also contained regions of the other OA manufacturing plasmids, indicating recombination between plasmid sequences (**Figure 4, Table 2**). Most of the complex structures and recombination involved the region between the ITRs of pSMN, the rep/cap region of pAAV2/9, and the region of pHelper containing the HAdV-derived genes (**Figure 4, Table 2**).

**Figure 4:**
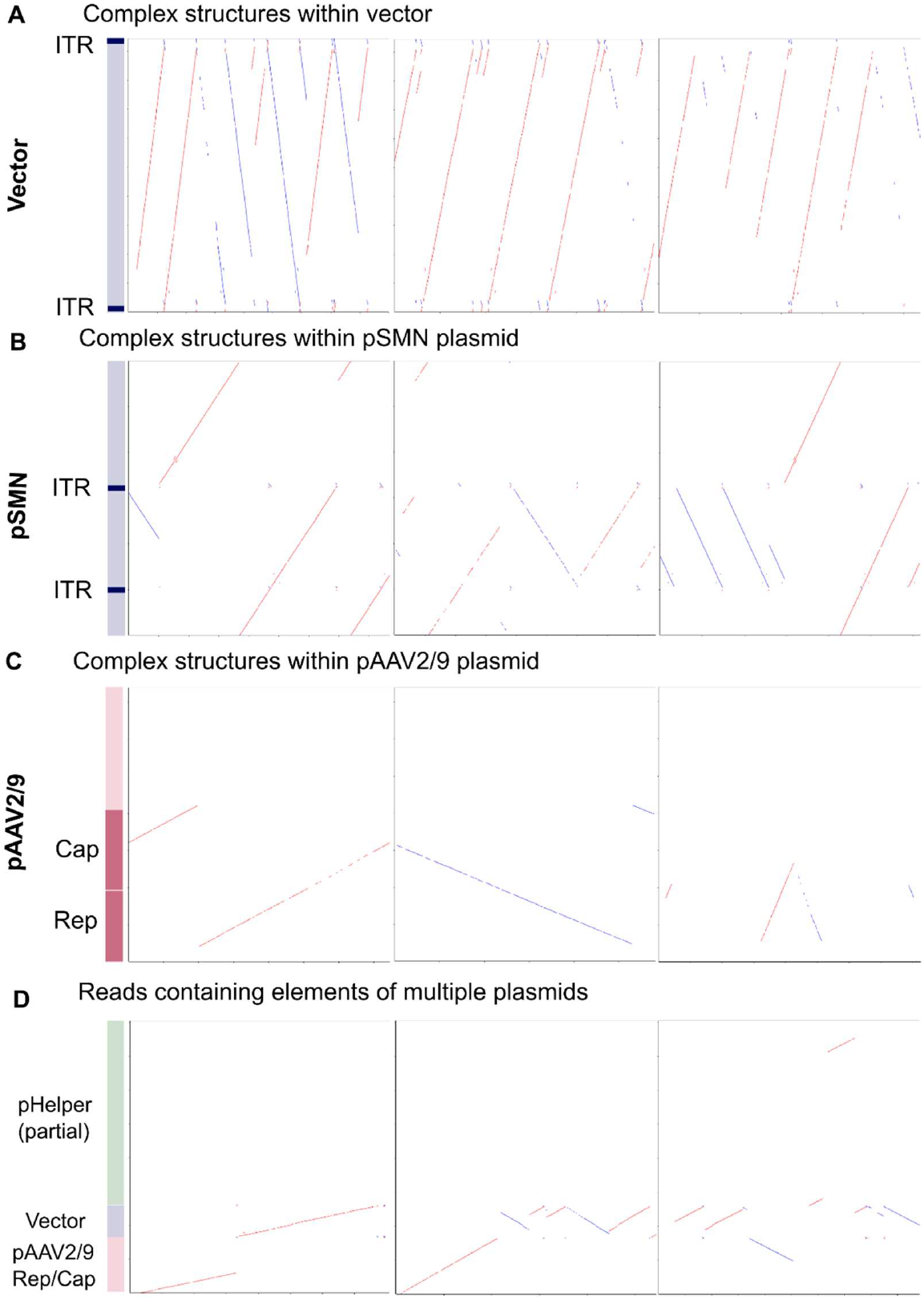
Alignment dot plots. Alignment dot plots showing individual nanopore reads (x axis) aligning to representative sequences of the OA manufacturing plasmids (y axis). Red dots show alignment to forward strand and blue to the reverse. A) Alignment against vector region of pSMN plasmid. B) Alignment against entire pSMN plasmid. C) Alignment to pAAV2/9 plasmid. D) Alignment to regions of all three plasmids – the vector region of pSMN, AAV rep and cap within pAAV2/9 and the HAdV gene region within pHelper. Representative images selected – numbers of reads belonging to each category can be found in Table 2.

**Table 2.** Gene therapy manufacturing plasmids in nanopore sequencing.

**2A**
| Region | Total reads | Total reads N50 | Filtered reads <sup>a</sup> | Filtered reads N50 | Filtered reads showing evidence of complex structures <sup>b</sup> |
| --- | --- | --- | --- | --- | --- |
| pSMN plasmid | 1533 | 10031 | 796 | 9244 | 683 |
| Vector only | 1517 | 10031 | 725 | 9103 | 611 |

**2B**
| Plasmid | Total reads | Total reads N50 | Region of interest (ROI) <sup>c</sup> | ROI reads | ROI reads N50 | ROI reads aligning to other plasmids <sup>d</sup> |
| --- | --- | --- | --- | --- | --- | --- |
| pAAV2/9 | 263 | 11006 | rep/cap genes | 88 | 10292 | 60 |
| pHelper | 194 | 11082 | All other than ori | 17 | 10443 | 9 |

<sup>a</sup> Reads are filtered based the following requirements: at least 1000 bp in length, 80% of the read aligns to the plasmid across all distinct alignments, and has a continuous stretch of matches/mismatches without insertions or deletions of at least 100 bp. This filtering helps remove reads that do not align convincingly to the plasmid sequence but allows for complexity in alignments due to concatemeric structures.
<sup>b</sup> Complex structures were identified using alignment dot plots. Monomeric structures and duplexes which might have arisen due to self-complementary vectors were excluded.
<sup>c</sup> Regions of interest contain the most relevant genes for AAV replication – AAV *rep* and *cap* in the case of pAAV2/9 and the adenovirus genes in the case of pHelper. These regions also exclude regions such as the origin of replication, which may be shared between plasmids.
<sup>d</sup> As determined by manual inspection of alignment dot plots.

Complex AAV vector genome rearrangements with structural rearrangements and concatemers have previously been demonstrated in macaque liver after treatment with rAAVs^31,32^ and in human hepatocytes in a humanised mouse model^33^. Similar complex concatemeric structures have been noted in liver samples from children with hepatitis associated with wild-type AAV2 infection^27^. To our knowledge, this is the first metagenomic analysis of liver tissue from an rAAV-treated patient to comprehensively elucidate the vector concatemer structures and complex genome rearrangements, which also included some manufacturing plasmid sequences.

Both short and long read metagenomic sequencing approaches in this study provide evidence that sequences from all three manufacturing plasmids were present in the liver of a patient with hepatitis after treatment with OA. Long read sequencing revealed large quantities of complexed plasmid DNA and many reads contained elements of multiple plasmids. It is possible that recombination events between plasmids took place during the manufacturing process, resulting in transduction of the AAV *rep* and *cap* genes and the HAdV helper genes as contaminants in the therapeutic vector preparation. Theoretically, this could allow replication competence of the vector genome in liver cells, thus explaining the replication-associated vector genome complexes we observed. The contribution of these genomic structures to the pathogenesis of rAAV hepatitis, and the putative role of contaminating plasmids and their potential immunogenicity, remain to be determined. It would be important to ascertain whether these genomic structures are also present in rAAV-treated patients without hepatitis.

Due to insufficient liver material, we were unable to assess whether the rearranged vector genomes were chromosomally integrated or episomal. Random, low frequency integration of various rAAV vectors in patient tissue is now well recognised^34–37^. AAV integrants in complex concatemers containing mixtures of rearranged and truncated vector genomes have been demonstrated in liver tissue of non-human primates after intravenous administration of rAAV8 vectors^31^. The significance of HHV-6B in the liver is unclear from this single case description. HHV-6 can act as a “helper” virus in wild-type AAV2 replication, and its genome contains a homologue of the AAV *rep* gene^38^. HHV-6 has also been found in liver tissue in a proportion of children with hepatitis associated with wild type AAV2 infection, although also sometimes in controls^27,28^, and in children with acute liver failure of unknown cause^39,40^.

Overall, our metagenomic analysis of liver tissue from a patient with hepatitis after OA treatment has revealed extensive disruption and concatemerisation of vector genomes, alongside numerous contaminating manufacturing plasmid sequences, with evidence of complex genomic structures and recombination events. We also identified HHV-6B in the liver. We postulate that presence of certain manufacturing plasmid sequences or helper viruses may allow replication of the vector genome within cells, giving rise to complex concatemeric structures. Future work is needed to determine the frequency and pathological significance of complex DNA structures in patient liver cells after rAAV gene therapy, whether they are episomal or integrated into the host genome, and how this may relate to the known hepatotoxicity of rAAV gene therapies.

## Data Availability

Data produced in the present study are available upon reasonable request to the authors.

## Acknowledgements

SB, OMT and SM are funded by the National Institute for Health Research (NIHR) Blood and Transplant Research Unit for Genomics to Enhance Microbiology Screening (NIHR203338). LB is funded by the NIHR Great Ormond Street Biomedical Research Centre (BRC). JB receives funding from the NIHR UCL/UCLH BRC. JB is an NIHR Senior Investigator. The views expressed are those of the author(s) and not necessarily those of the NHS, the NIHR or the Department of Health.

RK is funded by LifeArc P2020-0008 and P2023-0011, Great Ormond Street Hospital Children Charity and Dravet Syndrome UK Charity V4720 and V4919 and Therapeutic Acceleration Support (TAS), UCL.

This work was supported by grants CRUSH MC_UU_00034/9 and Wellcome Trust 226141/Z/22/Z.

The support of the GOSH and UCLH/ Institute of Neurology BRC to the Dubowitz Neuromuscular Centre Biobank is gratefully acknowledged.

The authors thank the team of the Histology Research Service, University of Glasgow, for the excellent technical support.

## Declaration of competing interests

GB is PI of clinical trials Sponsored by Roche, Novartis, Sarepta, Pfizer, NS Pharma, Reveragen, Percheron, Biomarin, Scholar Rock, and has received speaker and/or consulting fees from Sarepta, PTC Therapeutics, Entrada Therapeutics, Pfizer, Biogen, Novartis Gene Therapies, Inc. (AveXis), and Roche, and grants from Sarepta, Roche and Novartis Gene Therapies. UCL has received funding from Sarepta, Roche, Pfizer, Italfarmaco, Santhera.

FM is the PI of the Novartis sponsored trials in which OA was studied in the UK, and is also involved in clinical trials sponsored by Biogen, Roche, Sarepta Therapeutics, Genethon, PTC therapeutics and Solid Bioscience. He has received consulting fees from Pfizer, Sarepta, Roche, Biogen, Novartis, Solid, Dyne Therapeutics, Entrada, PTC and Edgewise.

MS is the sub-I of the Novartis sponsored trials in which OA was studied in the UK, and is also involved in clinical trials sponsored by Biogen, Roche, Dyne. She has received consulting fees from Roche, Biogen and Novartis.

### Methods Ethics

The liver biopsy procedure was performed for diagnostic purposes. Residual material was analysed in this study with informed consent for additional research under the International Severe Acute Respiratory and Emerging Infection Consortium (ISARIC) WHO Clinical Characterisation Protocol UK (CCP-UK) (ISRCTN 66726260). Ethical approval for the ISARIC CCP-UK study was given by the South Central–Oxford Research Ethics Committee in England (13/SC/0149), the Scotland A Research Ethics Committee (20/SS/0028) and the WHO Ethics Review Committee (RPC571 and RPC572).

### Short read metagenomic sequencing

Untargeted Illumina metagenomic sequencing of the liver biopsy was carried out by the clinical metagenomics service at Great Ormond Street Hospital, according to the protocol previously described^27,41^. A negative control sample consisting human DNA and RNA spiked with positive controls (cowpox DNA and FCV RNA) was run in parallel. Viruses were identified from the metagenomics data using Kraken2^42^ and Bracken^43^ run through Taxprofiler^44^, as well as metaMix^45^.

Human-filtered reads from the metaMix pipeline (other than for alignment to pSMN, where raw reads were used) were aligned using Bowtie2^46^ in very sensitive mode (apart from WT AAV2, where the parameters -score-min L,0,-0.1 -N 0 -L 22 --mp 6,2 --rdg 5,3 --rfg 5,3 were used) to genome sequences of AAV2 (NC_001401), HHV-6B (NC_000898) and HAdV-C (NC_001405) obtained from RefSeq, as well as representative sequences of the plasmids used in OA manufacture (pSMN^47^, pAAV2/9^48^, pHelper (pHGTI-Adeno1)^49^. The sequence of the AMR gene region in the pSMN plasmid did not match what was observed in the patient, so this region was reconstructed using the long read sequencing data and the modified pSMN sequence was used in all alignments. PCR duplicates were removed from the resulting alignments using samtools markdup^50^ and alignments were plotted using a custom R script.

### Long read metagenomic sequencing

DNA from approximately 3 mg of liver was purified using the Qiagen DNeasy Blood & Tissue kit as per the manufacturer’s instructions. DNA was fragmented to an average size of 10kb using a Megaruptor 3 (Diagenode) to reach an optimal molar concentration for library preparation. Quality control was perform using a Femto Pulse System (Agilent Technologies) and a Qubit fluorometer (Invitrogen). Samples were prepared for nanopore sequencing using the ligation sequencing kit SQK-LSK110. DNA was sequenced on a PromethION using R9.4.1 flowcells (Oxford Nanopore Technologies). Samples were run for 72 hours.

All library preparation and sequencing were performed by the UCL Long Read Sequencing facility.

Reads were trimmed using porechop with an adaptor threshold of 85 and were mapped to the human genome (ensemble GRCh38 v107) using minimap2^51^ in map-ont mode. Unaligned reads were then aligned to the regions of the plasmids shown in the figures using minimap2, and the aligned reads extracted using samtools^50^. A custom R script was used to filter reads that were over 1000bp in length, had a total alignment length of at least 80% of the total read length across all alignments and had a continuous stretch of matches/mismatches with no insertions or deletions of at least 100bp. Alignment dot plots for these reads were created using redotable^52^ with a window size of 20.

Representative examples are shown in the figures. Viruses were identified from the metagenomics data using Kraken2 and Bracken run through Taxprofiler.

### RNAscope *in situ-*hybridization

Formalin-fixed paraffin-embedded liver sections were cut at 2-3 µm thickness and mounted on glass slides. According to manufacturer’s instructions, RNAscope was performed with protease treatment and simmering in target solution (product codes: 322360 and 322331, ACDBio) to detect the SMN gene (product code: 553631, ACDBio, RNAscope® Probe - Hs-SMN1-CDS - Homo sapiens survival of motor neuron 1 telomeric (SMN1) transcript variant d mRNA). As positive control, a probe detecting Ubiquitin (product code: 310041, ACDBio) and as a negative control, a probe for DapB (product code: 310043, ACDBio) were used. Haematoxylin was used as a counterstaining and slides were digitised using the Leica Aperio 8 slide scanner.

### Immunohistochemistry

Immunohistochemistry was performed on formalin-fixed paraffin-embedded tissue cut at a thickness of 3 µm, using the Ventana Benchmark ULTRA staining platform and Optiview DAB Detection kit. The positive control was tonsil. The following antibodies were used: anti-CD4 (clone SP35, Roche, 790-4423), anti-CD8 (clone SP239, Roche, 790-7176) and anti-CD20 (clone L26, Dako (Agilent), M0755).

For all three antibodies, a HIER pre-treatment and a haematoxylin counterstain has been used.

## References

1. Blair, H. A. Valoctocogene Roxaparvovec: First Approval. Drugs 82, 1505–1510 (2022).

2. Heo, Y.-A. Etranacogene Dezaparvovec: First Approval. Drugs 83, 347–352 (2023).

3. Dhillon, S. Fidanacogene Elaparvovec: First Approval. Drugs 84, 479–486 (2024).

4. Hoy, S. M. Delandistrogene Moxeparvovec: First Approval. Drugs 83, 1323–1329 (2023).

5. Hoy, S. M. Onasemnogene Abeparvovec: First Global Approval. Drugs 79, 1255–1262 (2019).

6. Center for Biologics Evaluation and Research. Approved Cellular and Gene Therapy Products. FDA https://www.fda.gov/vaccines-blood-biologics/cellular-gene-therapy-products/approved-cellular-and-gene-therapy-products (2025).

7. Whiteley, L. O. An Overview of Nonclinical and Clinical Liver Toxicity Associated With AAV Gene Therapy. Toxicol Pathol 1926233231201408 (2023) doi:10.1177/01926233231201408.

8. Shieh, P. B., Kuntz, N. L., Dowling, J. J., Müller-Felber, W., Bönnemann, C. G., Seferian, A. M., et al. Safety and efficacy of gene replacement therapy for X-linked myotubular myopathy (ASPIRO): a multinational, open-label, dose-escalation trial. The Lancet Neurology 22, 1125–1139 (2023).

9. Mullard, A. Gene therapy community grapples with toxicity issues, as pipeline matures. Nature Reviews Drug Discovery 20, 804–805 (2021).

10. Chand, D., Mohr, F., McMillan, H., Tukov, F. F., Montgomery, K., Kleyn, A., et al. Hepatotoxicity following administration of onasemnogene abeparvovec (AVXS-101) for the treatment of spinal muscular atrophy. Journal of Hepatology 74, 560–566 (2021).

11. Finnegan, R., Manzur, A., Munot, P., Dhawan, A., Murugan, A., Majumdar, A., et al. Risk-benefit profile of onasemnogene abeparvovec in older and heavier children with spinal muscular atrophy type 1. Neuromuscul Disord 42, 22–26 (2024).

12. Gowda, V., Atherton, M., Murugan, A., Servais, L., Sheehan, J., Standing, E., et al. Efficacy and safety of onasemnogene abeparvovec in children with spinal muscular atrophy type 1: real-world evidence from 6 infusion centres in the United Kingdom. The Lancet Regional Health – Europe 37, (2024).

13. Shirley, J. L., Jong, Y. P. de, Terhorst, C. & Herzog, R. W. Immune Responses to Viral Gene Therapy Vectors. Molecular Therapy 28, 709–722 (2020).

14. Hösel, M., Broxtermann, M., Janicki, H., Esser, K., Arzberger, S., Hartmann, P., et al. Toll-like receptor 2–mediated innate immune response in human nonparenchymal liver cells toward adeno-associated viral vectors. Hepatology 55, 287–297 (2012).

15. Ashley, S. N., Somanathan, S., Giles, A. R. & Wilson, J. M. TLR9 signaling mediates adaptive immunity following systemic AAV gene therapy. Cellular immunology 346, 103997 (2019).

16. Larrey, D., Delire, B., Meunier, L., Zahhaf, A., de Martin, E. & Horsmans, Y. Drug-induced liver injury related to gene therapy: A new challenge to be managed. Liver Int (2024) doi:10.1111/liv.16065.

17. Bucher, K., Rodríguez-Bocanegra, E., Wissinger, B., Strasser, T., Clark, S. J., Birkenfeld, A. L., et al. Extra-viral DNA in adeno-associated viral vector preparations induces TLR9-dependent innate immune responses in human plasmacytoid dendritic cells. Sci Rep 13, 1890 (2023).

18. Hinderer, C., Katz, N., Buza, E. L., Dyer, C., Goode, T., Bell, P., et al. Severe Toxicity in Nonhuman Primates and Piglets Following High-Dose Intravenous Administration of an Adeno-Associated Virus Vector Expressing Human SMN. Human Gene Therapy 29, 285 (2018).

19. Audentes Therapeutics Inc (An Astellas Company). Comment on Docket FDA-2021-N-0651. https://www.regulations.gov/comment/FDA-2021-N-0651-0013.

20. Hordeaux, J., Lamontagne, R. J., Song, C., Buchlis, G., Dyer, C., Buza, E. L., et al. High-dose systemic adeno-associated virus vector administration causes liver and sinusoidal endothelial cell injury. Mol Ther 32, 952–968 (2024).

21. Groen, E. J. N., Talbot, K. & Gillingwater, T. H. Advances in therapy for spinal muscular atrophy: promises and challenges. Nat Rev Neurol 14, 214–224 (2018).

22. Zolgensma | European Medicines Agency (EMA). https://www.ema.europa.eu/en/medicines/human/EPAR/zolgensma (2020).

23. Wang, D., Tai, P. W. L. & Gao, G. Adeno-associated virus vector as a platform for gene therapy delivery. Nature reviews. Drug discovery 18, 358 (2019).

24. Srivastava, A., Mallela, K. M. G., Deorkar, N. & Brophy, G. Manufacturing Challenges and Rational Formulation Development for AAV Viral Vectors. J Pharm Sci 110, 2609–2624 (2021).

25. Brimble, M. A., Cheng, P.-H., Winston, S. M., Reeves, I. L., Souquette, A., Spence, Y., et al. Preventing packaging of translatable P5-associated DNA contaminants in recombinant AAV vector preps. Molecular Therapy Methods & Clinical Development 24, 280–291 (2022).

26. Center for Biologics Evaluation and Research. Chemistry, Manufacturing, and Control (CMC) Information for Human Gene Therapy Investigational New Drug Applications (INDs). https://www.fda.gov/regulatory-information/search-fda-guidance-documents/chemistry-manufacturing-and-control-cmc-information-human-gene-therapy-investigational-new-drug (2020).

27. Morfopoulou, S., Buddle, S., Torres Montaguth, O. E., Atkinson, L., Guerra-Assunção, J. A., Moradi Marjaneh, M., et al. Genomic investigations of unexplained acute hepatitis in children. Nature 617, 564–573 (2023).

28. Ho, A., Orton, R., Tayler, R., Asamaphan, P., Herder, V., Davis, C., et al. Adeno-associated virus 2 infection in children with non-A–E hepatitis. Nature 617, 555–563 (2023).

29. Chapin, C. A., Melin-Aldana, H., Kreiger, P. A., Burn, T., Neighbors, K., Taylor, S. A., et al. Activated CD8 T-cell Hepatitis in Children With Indeterminate Acute Liver Failure. Journal of Pediatric Gastroenterology and Nutrition 71, 713–719 (2020).

30. Meier, A. F., Tobler, K., Leisi, R., Lkharrazi, A., Ros, C. & Fraefel, C. Herpes simplex virus co-infection facilitates rolling circle replication of the adeno-associated virus genome. PLoS Pathog 17, e1009638 (2021).

31. Greig, J. A., Martins, K. M., Breton, C., Lamontagne, R. J., Zhu, Y., He, Z., et al. Integrated vector genomes may contribute to long-term expression in primate liver after AAV administration. Nat Biotechnol 42, 1232–1242 (2024).

32. Sun, X., Lu, Y., Bish, L. T., Calcedo, R., Wilson, J. M. & Gao, G. Molecular analysis of vector genome structures after liver transduction by conventional and self-complementary adeno-associated viral serotype vectors in murine and nonhuman primate models. Hum Gene Ther 21, 750–61 (2010).

33. Dalwadi, D. A., Calabria, A., Tiyaboonchai, A., Posey, J., Naugler, W. E., Montini, E., et al. AAV integration in human hepatocytes. Molecular Therapy 29, 2898–2909 (2021).

34. Gil-Farina, I., Fronza, R., Kaeppel, C., Lopez-Franco, E., Ferreira, V., D’Avola, D., et al. Recombinant AAV Integration Is Not Associated With Hepatic Genotoxicity in Nonhuman Primates and Patients. Molecular Therapy 24, 1100–1105 (2016).

35. Kaeppel, C., Beattie, S., Fronza, R., Logtenstein, R., Salmon, F., Schmidt, S., et al. A largely random AAV integration profile after LPLD gene therapy. Nature medicine 19, (2013).

36. Schmidt, M., Foster, G. R., Coppens, M., Thomsen, H., Dolmetsch, R., Heijink, L., et al. Molecular evaluation and vector integration analysis of HCC complicating AAV gene therapy for hemophilia B. Blood Advances 7, 4966–4969 (2023).

37. Symington, E., Rangarajan, S., Lester, W., Madan, B., Pierce, G. F., Raheja, P., et al. Long-term safety and efficacy outcomes of valoctocogene roxaparvovec gene transfer up to 6 years post-treatment. Haemophilia 30, 320–330 (2024).

38. Thomson, B. J., Weindler, F. W., Gray, D., Schwaab, V. & Heilbronn, R. Human Herpesvirus 6 (HHV-6) Is a Helper Virus for Adeno-Associated Virus Type 2 (AAV-2) and the AAV-2 rep Gene Homologue in HHV-6 Can Mediate AAV-2 DNA Replication and Regulate Gene Expression. Virology 204, 304–311 (1994).

39. Warner, S., Brown, R. M., Reynolds, G. M., Stamataki, Z. & Kelly, D. A. Case report: Acute liver failure in children and the human herpes virus 6-? A factor in the recent epidemic. Front Pediatr 11, 1143051 (2023).

40. Yang, C. H., Sahoo, M. K., Fitzpatrick, M., Lau, A. H., Pinsky, B. A. & Martinez, O. M. Evaluating for Human Herpesvirus 6 in the Liver Explants of Children With Liver Failure of Unknown Etiology. The Journal of Infectious Diseases 220, 361–369 (2019).

41. Atkinson, L., Lee, J. CD., Lennon, A., Shah, D., Storey, N., Morfopoulou, S., et al. Untargeted metagenomics protocol for the diagnosis of infection from CSF and tissue from sterile sites. Heliyon 9, e19854 (2023).

42. Wood, D. E., Lu, J. & Langmead, B. Improved metagenomic analysis with Kraken 2. Genome Biology 20, 257 (2019).

43. Lu, J., Breitwieser, F. P., Thielen, P. & Salzberg, S. L. Bracken: estimating species abundance in metagenomics data. PeerJ Comput. Sci. 3, e104 (2017).

44. taxprofiler: Introduction. https://nf-co.re/taxprofiler/1.0.1.html.

45. Morfopoulou, S. & Plagnol, V. Bayesian mixture analysis for metagenomic community profiling. Bioinformatics 31, 2930–2938 (2015).

46. Langmead, B. & Salzberg, S. L. Fast gapped-read alignment with Bowtie 2. Nat Methods 9, 357–359 (2012).

47. Kaspar, B. K., Burghes, A. & Porensky, P. Intrathecal delivery of recombinant Adeno-associated virus 9. (2022).

48. Gao, G., Wilson, J. & Alvira, M. Adeno-associated virus (aav) serotype 9 sequences, vectors containing same, and uses therefor. (2005).

49. Gray, J. Molecule Information, pHGTI-Adeno1, Harvard Gene Therapy Initiative. (2004).

50. Li, H., Handsaker, B., Wysoker, A., Fennell, T., Ruan, J., Homer, N., et al. The Sequence Alignment/Map format and SAMtools. Bioinformatics 25, 2078 (2009).

51. Li, H. Minimap2: pairwise alignment for nucleotide sequences. Bioinformatics 34, 3094–3100 (2018).

52. Andrews, S. s-andrews/redotable. (2024).

